# Prognostic Value of CD8+ T-Cells at the Invasive Margin is Comparable to Immune Score in Non-Metastatic Colorectal Cancer: Prospective Multicenter Cohort Study

**DOI:** 10.1101/2024.09.23.24314210

**Authors:** Durgesh Wankhede, Niels Halama, Matthias Kloor, Dominic Edelmann, Hermann Brenner, Michael Hoffmeister

## Abstract

**Background:** The Immunoscore® is a validated tool for predicting colorectal cancer (CRC) prognosis, yet its adoption is impeded by complex commercial software and patient reimbursement challenges. Utilizing open-source methods, this study aimed to explore whether an immune cell score can be facilitated by focusing on single T-cell markers, to provide a simplified prognostic model in non-metastatic CRC.

**Methods:** A multicentric prospective cohort study was conducted in non-metastatic CRC patients who underwent curative surgical resection. CD3+ and CD8+ tumor infiltrating lymphocytes (TILs) were quantified in both invasive margin (IM) and tumor core (TC) using QuPath. A composite score, termed immune cell score, mirroring the methods employed for the Immunoscore®, was calculated based on the TIL densities (CD3-IM, CD8-IM, CD3-TC, CD8-TC]. We used a split sample approach (70:30) to estimate adjusted hazard ratios of cancer-specific survival (CSS) in a training and a validation set. Classification and regression tree analysis (CART) was performed to select the most prognostic TIL. The model incorporating the CART-selected TIL was compared to a two-tiered immune cell score model for overall performance (Brier score) and discrimination (concordance probability estimate, CPE).

**Results:** During a median follow-up time of 9.0 years, among 1260 patients, there were 203 CRC specific deaths. CART-selected CD8-IM was the most prognostic TIL at a cut-off of 231 cells/mm2. Patients with CD8-IM^Hi^ had better CSS than CD8-IM^Low^ in both training (HR 0.58, 95% CI 0.40-0.84) and validation sets (HR 0.35, 95% CI 0.21-0.60). Brier scores of CD-8IM and immune cell score survival models were comparable in both training and validation cohort, whereas the survival discrimination of CD8-IM slightly outperformed the immune cell score in the validation set (CPE: CD8-IM 0.748, IS 0.730).

**Conclusion:** A single TIL marker, specifically CD8-IM, provided prognostic information comparable to the immune cell score. Simplified and cost-effective TIL assessments could enhance their bench to bedside translation and may guide adjuvant therapy in early-stage CRC.

## Introduction

Colorectal cancer (CRC) ranks second in cancer-specific deaths worldwide and is the primary cause of cancer death among younger men (< 50 years)^1^. In addition to the increase in early-onset CRC over the last decade, there is a rising trend in the incidence of regional stage CRC [stage III; 2%-3% annual percent change]^2^. Up to 50% of stage III colon cancer patients experience recurrence following curative resection, often requiring adjuvant chemotherapy ^3^. In stage III colon cancer patients, the TNM staging is less effective in differentiating prognostic groups [5-year cancer-specific survival (CSS): 37% - 67%], especially among those who underwent adjuvant chemotherapy^4^. Despite variable clinical outcomes within the same stage, the stage at diagnosis remains the cornerstone in CRC prognostication and therapy decisions, thus indicating an urgent need for a more robust risk-stratification system^5^.

Tumor-infiltrating lymphocytes (TILs) reflect an interplay between the tumor and adaptive immune system and are an integral component of the tumor microenvironment ^6^. Multiple studies have shown that higher TIL densities in primary tumors is associated with favorable survival outcomes^7-9^. CD3+, CD8+, and FOXP3+ T-cells exert dominant prognostic effects in CRC, even after adjusting for pathologic stage and microsatellite instability (MSI) status ^8,10^. The spatial influence of TILs in the tumor microenvironment is evident, as TILs in tumor core (TC) and invasive margin (IM) showed independent associations with CRC survival outcomes^11^. Evidence from clinical trials suggests a role of immune cell densities in optimal risk stratification of stage III colon cancer patients for adjuvant chemotherapy, consolidating their role as a promising predictive biomarker^7,12^.

TIL assessment, characterization, and quantification methods have evolved from manual counting by pathologists under light microscope to automatic annotation and counting using digital pathology applications^13,14^. One of the prominent prognostic assays in CRC is the “Immunoscore®” which is based on CD3+ and CD8+ T-cell quantification in TC and IM ^12,15^. The mean percentiles of the T-cells densities (CD3-IM, CD8-IM, CD3-TC, & CD8-TC) are converted into combined immune marker score using a commercial software^15^. Despite the multicentric validation of Immunoscore, its widespread clinical adoption is limited by the reliance on commercial software and the need for the patients to either pay out of pocket or reimbursement by the insurance providers^16,17^. We previously validated the prognostic potential of the Immunoscore in CRC patients by employing open-source software and applying comparable methods for immune cell analysis^18^.

A simplified approach focusing on single TILs might offer comparable prognostic information to the Immunoscore, and could facilitate its bench-to-beside translation. In this study, we analyzed individual components of Immunoscore (CD3-IM, CD8-IM, CD3-TC, and CD8-TC) on resected tumor specimens for their prognostic association in a large population-based nonmetastatic CRC cohort, using an open-source digital pathology application. Intending to deliver a more simplified Immunoscore, we employed a decision tree algorithm used in machine learning to select the most prognostic immune marker and compared it with the Immunoscore model for cancer-specific survival (CSS).

## Methods

### Study population

The “Darmkrebs: Chancen der Verhütung durch Screening” (DACHS) study is a population-based case-control study on CRC, recruiting participants from southwest Germany with long-term follow-up of the patients conducted at 3, 5, and 10 years after diagnosis^19^. Patients in the present study were recruited between 2003 and 2010 and had the following inclusion criteria: a) stage I-III CRC (stage updated based on the 8^th^ American Joint Committee on Cancer TNM system^5^); b) curative resection (R0); c) had not undergone neoadjuvant therapy (chemotherapy &/or radiotherapy); d) availability of tumor formalin-fixed, paraffin-embedded (FFPE) blocks with both CD3+ and CD8+ immunohistochemistry (IHC) stains (**Figure 1A**). Each participant provided written informed consent at inclusion into the study. The research ethics committee of the medical faculty at University of Heidelberg and the state medical boards of the Baden-Wuerttemberg and Rhine-Palatinate approved the present study (Ethical approval number: 310/2001). We adhered to the guidelines outlined in the Reporting recommendations for tumour MARKer prognostic studies (REMARK) (**Table S1**)^20^.

**Figure 1.** Overview of the study protocol. A. Immune cells quantification using a machine learning digital pathology platform; B. Deriving immune cell score from individual immune cell density; C. Classification and Regression Tree (CART) analysis to determine the most prognostic immune cell.

### Immunohistochemistry and digital image analysis

TIL densities and IS assessment were performed through whole-section image analysis using an open-source digital pathology application, as previously described^18^. In brief, two consecutive tumor FFPE block sections were stained with rabbit monoclonal antibodies for CD3+ and CD8+ T-cells evaluation (2GV6 & SP57 respectively, Roche Diagnostics) using the Ventana BenchMark Ultra Slide Stainer (Roche Diagnostics). The Aperio AT2 slide scanner digitized the stained slides (Leica Biosystems, Germany) at a 40x magnification (0.25 μm/pixel) following manufacturer’s protocols. Each scanned slides were analyzed with QuPath software version 0.5.0, with manual annotation of tumor borders^21^. The 1-mm edge of the tumor where the invasive tumor gland infiltrates the surrounding normal tissue (500 μm in the tumor area and 500 μm in the normal tissue) was defined as IM, whereas the rest of the tumor was considered as TC. Punch areas in the FFPE blocks from previous sampling, artifacts, normal tissue, and necrosis were demarcated and not considered.

We used a previously validated immune cell score as the benchmark, derived from methods described by the international Immunoscore consortium^15,18^. An automated positive cell detection algorithm from the software was employed with the optimized thresholds for CD3+ and CD8+ T-cells (**Table S2**). Positive cell densities in the two regions were calculated as the number of T-cells per square millimeter. T-cell densities (CD3-IM, CD3-TC, CD8-IM, CD3-TC) were converted to percentiles. The mean of these percentiles yielded a two-level immune cell score (low (ICS^Lo^), high (ICS^Hi^)) and a three-level immune cell score (low, intermediate (ICS^Int^), high). For the two-level immune cell score, a 25th percentile cutoff was used, and for the three-level system, cutoffs at 25th and 70th percentiles were applied (**Figure 1B**)^12,15,18^. The methodologies employed for assessing MSI status and *KRAS* and *BRAF*^*V600E*^ mutations were previously outlined^18,22^.

### Statistical analysis

The primary endpoint of the study was CSS, measure from the time of surgery until CRC-related death. Quartiles [Q1 to Q4] of TIL densities were employed to investigate the relationship between each immune marker and CSS, with Q1 serving as the reference category. Kaplan-Meier plots and the log-rank test were used to determine the survival differences between quartiles.

To investigate a more simplified prognostic model, the cohort was randomly divided into training (n = 882, 70%) and validation set (n = 378, 30%). Two statistical approaches were used to build a simplified TIL models. The first approach used classification and regression tree analysis (CART) on the training set to select the most prognostic TILs and determine optimal cut-off values for dichotomous categorization (**Figure 1C**)^23^. CART, a machine learning method, identifies variance patterns in CSS relative to TIL densities, accommodating both linear and non-linear relationships without data transformation. Dichotomous splitting continued until the terminal leaf had 20 observations. A 5-fold cross-validation (80% training, 20% validation) with the lowest cross-validation error pruned the CART model. The terminal groups from the pruned model were used as categorical variables in survival analyses. Random survival forests assessed the stability of CART results^24^. This ensemble method builds multiple decision trees to investigate right-censored survival data, combining their predictions to improve performance^25^. Variable importance was calculated using an aggregate of 1000 trees including all four immune markers^26^. Agreement between CART and random survival forest results would corroborate findings.

The association of the prognostic CART-derived TIL group with CSS was assessed in the training and validation set using a multivariable Cox regression model, adjusting for covariates including age, sex, T/N stage, tumor side, histologic grade, adjuvant chemotherapy, MSI status, *KRAS*, and *BRAF* mutations. The association of immune cell score with CSS was determined adjusting for same covariates. The covariates adjusted in the multivariable Cox models were age, T/N stage, tumor side, adjuvant chemotherapy, *BRAF* mutation, sex, histologic grade, MSI status, and *KRAS* mutation^27,28^.

The second approach involved applying cut-off values corresponding to the 25th and 70th percentiles of TIL density, mirroring those of the Immunoscore, to each immune marker in both the training and validation sets. Multivariable Cox regression models were constructed with each immune marker treated as a categorical variable and compared against the immune cell score model. Associations with CSS were reported as adjusted hazard ratios (HR_adj_) and 95% confidence intervals (CIs).

Model performance derived from both approaches were determined using an integrated Brier score (range, 0-1), with lower scores indicating superior accuracy of the probabilistic survival predictions^26^. Discriminatory power was assessed using the concordance probability estimate (CPE), indicating the likelihood that a patient who had the better outcome from the Cox model will a longer survival ^29^. Over-optimism of models trained on the training set was assessed using a bootstrap resampling approach^30^. Statistical significance was determined using two-sided *p*-values, and results with *p*-values less than 0.05 were considered significant. R-Studio version 4.2.2 was used to perform the statistical analyses.

## Results

After quality control, 1260 stage I-III CRC patients were included in the study. Of these, 57.6% were men and the median age was 71 years (interquartile range (IQR) 63-78 years). An MSI genotype was observed in 14.7% of the total cohort. The median follow-up was 9 years (IQR, 5-10) during which 203 patients died due to CRC. The patient characteristics are presented in **Table S3**.

### TILs and Clinical Parameters

CD3+ T-cell densities were greater than those of CD8+ T-cells, and IM densities exceeded those of the TC (**Figure S1**; p < 0.0001). CD3-IM exhibited the highest median density at 1134 cells/mm^2^ (IQR: 710-1686), while CD8-TC demonstrated the lowest density at 219 cells/mm^2^ (IQR: 122-436) (**Figure S2**; Shapiro-Wilk test p <0.0001). Both CD3+ and CD8+ T-cells showed significant positive correlation in both IM and TC (r > 0.6, p <0.05) (**Figure S3**). Right-sided tumors had higher TIL densities except for CD3-TC, compared to the left side (p <0.001) (**eFigure 4A**). Lower pathological T and N stages were associated with higher densities of all immune markers (**eFigure 4B-C**). No association was observed between TIL densities and the number of involved nodes post-nodal metastasis. Patients receiving adjuvant chemotherapy had lower T-cell densities than chemotherapy-naïve patients (**eFigure 4D**). MSI and *BRAF*^*V600E*^ mutated tumors showed significantly higher T-cell densities than MSS and *BRAF* wild-type, respectively (**eFigure 4E-F**). CD3+ densities in the TC were significantly higher in *KRAS*-wild type than in mutated tumors (**eFigure 4G**). Age, sex, and tumor grade did not show an association with any of the T-cell densities.

### Associations of TILs with Cancer-Specific Survival

#### TIL Densities as Quartiles

In the overall cohort, survival analyses by modeling the T-cell densities as quartiles demonstrated that patients with the highest densities (Q4) for all four immune markers exhibited favorable survival, compared to lower densities (log-rank tests, *p* < 0.0001) (**eFigure 5A-D**).

#### CART model

In the training set, the unpruned CART model identified CD8-IM as the primary prognostic immune marker, setting a cut-off at 231 cells/mm^2^ (22%). CD3-IM was the second most prognostic marker, with a cut-off of 309 cells/mm^2^. Through 5-fold cross-validation, CD8-IM emerged as the most prognostic marker in three folds, CD3-IM in one, and CD8-IM/CD3-IM in the last fold. The pruned CART model, with the lowest cross-validation error (complexity parameter = 0.02), selected CD8-IM as the most prognostic TIL (**Figure 2**). CD8-IM was also selected as the variable of important through the random survival forest, with the lowest out-of-the-bag error rate of 0.41, further corroborating the CART results (e**Figure 6**).

**Figure 2.** Pruned Classification and Regression Tree from the training cohort with CD8IM as the solely selected variable and cancer-specific survival as the outcome. The numbers in the node from top of bottom: survival probability, number of events/number of observations, percentage observations in the node.

Based on the pruned CART model, CD8-IM was categorized into high (CD8-IM^Hi^) and low (CD8-IM^Lo^) subgroups with 231 cells/mm^2^ as the cut-off value. In the training set, the 5-year survival of for CD8-IM^Hi^ and CD8-IM^Lo^ was 89% (95% CI 87-91.7%) and 77% (95% CI 71-84%), respectively. In the validation set, 5-year survival for CD8-IM^Hi^ was 92% (95% CI 89-95%) and 79% (95% CI 71-88%) in the CD8-IM^Lo^ subgroup (**Figure 3A-D**). In a multivariable Cox model, patients with CD8-IM^Hi^ had significantly better CSS than patients with CD8-IM^Lo^ in the training (HR_adj_ 0.58, 95% CI 0.40-0.84) and validation sets (HR_adj_ 0.35, 95% CI 0.21-0.60]. The HR_adj_ for ICS^Hi^ was 0.77 (95% CI 0.55-1) and 0.68 (95% CI 0.39-1.1) in the training and validation sets, respectively (**Figure 4**).

**Figure 3.** Kaplan-Meier analysis of cancer-specific survival of immune markers. A. Training cohort: CD8IM, B. Training cohort: immune cell score, C. Validation cohort: CD8IM, D. Validation cohort: immune cell score

**Figure 4.** Classification and regression tree selected immune marker versus immune cell score. A. Sankey diagram showing the distribution of patients in the immune marker subgroup**s** for CD8IM and immune cell score, B. Multivariable Cox regression analysis for CART selected immune marker and immune cell score for cancer-specific survival. ^*^Adjusted for age, sex, tumor and nodal stage, adjuvant chemotherapy, tumor grade, microsatellite instability status (MSI), *KRAS* and *BRAF* mutation. ^Cutoff value = 231 cells/mm^2. $^Cutoff value = 25 mean percentile immune cell density

#### Standard Cut-off Values

The CD3-IM group demonstrated identical patient distribution, event occurrences, and outcomes in both training and validation sets when compared to the immune cell score group, indicating that the results obtained using standard cut-off values for CD3-IM accurately reflects the immune cell score group. In the training set, all four immune markers exhibited favorable survival in the high subgroups compared to the low subgroups. CD8-TC had the strongest association with CSS [HR_adj_ 0.45, 95% CI 0.27-0.75]. CD3-TC [HR_adj_ 0.66, 95% CI 0.41-1.08] showed an association in the same direction, though less robust. For CD8-IM, compared to the low subgroup, patients in high subgroup showed strong associations with longer CSS [HR_adj_ 0.51, 95% CI 0.30-0.87], whereas those in the intermediate subgroup, the association was slightly less robust (HR_adj_ 0.77, 95% CI 0.53-1.12). The HR_adj_ for the ICS^Int^ and ICS^Hi^ subgroups were 0.90 (95% CI 0.62-1.32) and 0.51 (95% CI 0.31-0.87), respectively (**Figure 5**).

**Figure 5.** Individual immune markers versus immune cell score: Three level classification. 1. CD3IM vs immune cell score, A. Sankey diagram, B. Multivariable Cox regression analysis for CD3IM and immune cell score. 2. CD8IM vs immune cell score, A. Sankey diagram, B. Multivariable Cox regression analysis for CD8IM and immune cell score. 3. CD3TC vs immune cell score, A. Sankey diagram, B. Multivariable Cox regression analysis for CD3TC and immune cell score. 4. CD8TC vs immune cell score, A. Sankey diagram, B. Multivariable Cox regression analysis for CD8TC and immune cell score. ^*^Adjusted for age, sex, tumor and nodal stage, adjuvant chemotherapy, tumor grade, microsatellite instability status (MSI), *KRAS* and *BRAF* mutation. ^a^Cutoff value = 25 percentile immune cell density. ^^^Cutoff value = 70 percentile immune cell density

In the validation set, the intermediate and high subgroups of all immune markers showed a trend towards improved survival, though the ranges suggested variable precision. CD8-IM demonstrated a more pronounced association in the high subgroup (HR_adj_ 0.40, 95% CI 0.22-0.73) and modest association in the intermediate subgroup (HR_adj_ 0.49, 95% CI 0.23-1.02). The HR_adj_ of 0.70 (95%CI 0.38-1.29) for ICS^Int^ and 0.64 (95% CI 0.31-1.29) for ICS^Hi^ subgroups suggested a positive association of immune cell score with CSS (**Figure 5**).

### Discrimination ability and Overall Performance of Survival Models

Applying CART analysis on the training set, the CPE for the CD8-IM and immune cell score Cox models were 0.722 and 0.728, suggesting comparable discriminative ability. A slightly improved discriminative ability of the CD8-IM model (CPE 0.738) was observed compared to the immune cell score model (CPE 0.726) in the validation set, which was retained after optimism correction (CPE_adj_: CD8-IM 0.748; ICS 0.730).

When employing standard cut-off values, the CPE obtained from the Cox models for all immune markers demonstrated comparable survival discrimination, both among each other (range 0.725 -0.729) and even to immune cell score (0.729) in the training set. In the validation set, the CPE adjusted for over-optimism from CD8-IM (0.747) exhibited slightly superior discrimination compared to other immune markers and even outperformed the immune cell score (0.736) (**Table S3**).

Both the CART derived CD8-IM model and the two-level immune cell score model exhibited comparable overall performances in both training and validation sets over 10 years of follow-up. The three-tiered immune marker models had similar performance to the three-level immune cell score model in both training and validation sets (**Table S4**).

## Discussion

In the present study, we utilized transparent TIL assessment methods using open-source digital pathology software and compared the prognostic value of CD3-IM, CD3-TC, CD8-IM, and CD8-TC to the immune cell score. Using CART analysis and incorporating these four TILs as continuous variables, CD8-IM was selected as the most prognostic immune marker. In the two-tiered survival models, CD8-IM was an independent predictor of CSS, and showed comparable performance to immune cell score. On using the standard cut-off values [25% and 70%] for TIL densities, the high-density subgroups of all four immune markers exhibited favorable survival compared to the low subgroups. Notably, these three-tiered survival models of all four immune markers demonstrated similar performance to the immune cell score model in the training cohort, and CD8IM moderately outperformed the rest in the validation cohort. These results could be of significant clinical importance considering that intra-immune cell and spatiotemporal heterogeneity deter the translation of TILs in routine clinical practice^31^.

### Comparison with Previous Studies

Apart from combined TIL performance, the single markers were found to provide additional prognostic information, reflecting a comprehensive overview of tumor immune-microenvironment^6^. Previous studies showed a positive association of CD3+ TILs with survival in CRC; however, with certain caveats^32^. CD3 is a highly specific IHC marker for T-cells; however, studies have shown that CD3 expression levels differ across various T-cell populations, indicating distinct phenotypes and potentially different functional properties within the immune system^33^. High-dimensional cytometry and RNA-based single-cell analysis revealed heterogeneity in the CD3+ T-cell population^34^. On the other hand, although CD8+ TILs also show diverse functional profiles, they consistently show stronger associations with better survival in CRC than CD3+ TILs, which also aligns with our results^35^. To differentiate between cancer cells-reactive CD8+ TILs to their bystander variants, a few studies utilized labeling activation markers, such as SDF-1, and TNF-α, and effector molecules like perforin and granzymes B^36-38^. In these studies, activation markers either demonstrated independent prognostic value or, when combined with CD8+ expression, exhibited a stronger association with survival compared to high levels of CD8+ expression alone. Considering the prominent role of CD8+ T-cells in the function of immune checkpoint inhibitors, activated CD8+ TILs may provide more informative insights into the tumor immune milieu than other immune markers, and thus serve as a better biomarker than isolated CD8+ expression^39^.

In addition to TIL subtypes, Pages et al. suggested an improved survival prediction with combined analysis of TC and IM, compared to single location analysis^40^. Interestingly, in addition to CD8+ T-cells, CD45RO+ instead of CD3+ T-cells were assessed in this study. Survival varied by TIL subtypes and location, with high TIL density in both tumor regions showing the best survival compared to uniformly low TIL density. However, in the heterogenous subgroup, patients with TC^Hi^-IM^Low^ TILs were combined with TC^Low^-IM^Hi^, which restricted the prognostic interpretation of individual locations^40^. Similarly, a seminal proof of concept study revealed potentially favorable survival in patients with high TIL density in combined regions compared to low TIL density^41^. CD3+ and CD45RO+, instead of CD8+ TILs, were evaluated in this study and it remains unclear if the combined TIL subtype and location provided additional prognostic value compared to the individual TIL subtypes and locations.

### Clinical Implications and Future Directions

Fulfilment of three assumptions, namely, clinical validity, analytical validity, and clinical utility, are necessary for a tumor biomarker to be translated from bench to bedside^42^. In this context, the Immunoscore and its individual components have been clinically validated to predict OS, DFS, and CSS in both localized and metastatic colorectal cancer^13,15,18,32,39,43^. Furthermore, we previously validated the Immunoscore using open-source digital pathology application by adapting an analogous scoring method in a large CRC cohort, which also served as the comparative benchmark in the present study^18^. Despite TIL density being a continuous variable, a threshold is often sought for clinical decision, which could be derived from statistically driven methods to arbitrary cut-off values. In the international Immunoscore® consortium study, the lower (25%) and upper cut-off values (70%) were selected for a balanced distribution of patients in each subgroup, aiming for approximately a quarter in both low and high categories; albeit, the statistical relevance of these thresholds to the outcome was not evaluated^15^. In the present study, the CART analysis allowed to integrate TILs as continuous variables, preserving their statistical power and relevance^44^. Nonetheless, prospective biomarker studies are essential to obtain a generalizable cut-off value for TILs for their successful clinical translation.

In the present study, survival predictions of TILs at the IM were consistently stronger than at the TC for both CD3 and CD8, which corroborates with current evidence^45^. Prognostic superiority of TILs at IM is further highlighted by the crucial interplay between pro- and anti-tumor factors at the tumor-normal tissue interface, where TILs may prevent cancer cell dissemination^46^. Findings from our study support this contention, showing higher TIL density in tumors without nodal metastasis or deeper local invasion. Moreover, studies have demonstrated that tumors with low TIL density exhibited strong association with high-risk pathological factors for metastatic invasion (venous embolism, lymphatic and perineural invasion) and epithelial-to-mesenchymal transition^10,11,47^. We also found TILs at IM had a two-fold higher median cell count than at the TC. Evidence suggests TILs at the TC may not adequately reflect anti-tumor immune status due to their limited presence in this region compared to the IM, reducing their clinical utility despite their clinical validity^48^.

A recent study has shown that the IHC antibodies, DAB kits, operators, and instruments do not influence staining of TILs and prognostic classification of the Immunoscore^49^. Furthermore, TIL staining intensities remained unaffected by age of tumor blocks or position of tissue sections, consolidating its analytical validation^49^. The methods employed in our analysis mirrored those of the international Immunoscore® consortium^15^ (**Table S5**). Nonetheless, certain differences prevailed, such as width of IM (Immunoscore: 760 µm, immune cell score: 1000 µm), and most importantly, the digital pathology platform. Immunoscore Analyzer (Veracyte SAS, Marseille, France) is employed to derive the Immunoscore^50^. However, as it is a closed-sourced platform, the coding and detailed assessment methods are not publicly accessible. We relied on QuPath, which is an open-source, cross-platform tool that uses machine learning techniques for biomarker quantification and WSI annotation and visualization^21^.

For clinical implementation, machine learning algorithms need rigorous evaluation to ensure their prognostic reliability. “Black Box” is a term used in the context of machine learning, where the derivation methods of the trained algorithms are often unknown^51^. More efforts are being made to increase transparency and convert this “black-box” to “glass-box” through “explainable” machine learning^52^. Open-source digital pathology applications offer accessible and customizable source codes based on the user’s requirements, unlike commercial software, which are designed for specific functions. Closed-source platforms can be prohibitively expensive for independent researchers, small laboratories, and those in resource-limited regions. In contrast, open-source tools offer affordability, consistency, and foster collaboration among pathologists. This accessibility can be particularly beneficial for remote or understaffed areas, especially during events such as the COVID-19 pandemic, potentially leading to decreased healthcare costs^53^.

### Limitations

One of the primary limitations of our study is that we did not use the original trademarked Immunoscore as the comparative benchmark. Although our previous research demonstrated that it was possible to implement an analogous immune cell score by adapting the methodology of the Immunoscore from published literature which yielded very similar results, this similarity is based on indirect validation rather than direct comparison^18^. While the study adjusts for several covariates, there might still be unmeasured confounders that could influence the observed associations. Due to inclusion of patients from a specific geographical region, generalizability of our findings may be limited. Lastly, although the models were validated using separate training and validation sets, external validation with independent cohorts could not be performed. Given the challenges in procuring independent cohort, the internal validation set, although derived from the same study population, serves as a widely utilized method to test the reproducibility of the findings. While the validation set is smaller, cross-validation within the CART analysis helps mitigate overfitting concerns. Furthermore, the use of random survival forests to assess the stability of the CART results and use of bootstrap analysis to correct over-optimism in the results from validation sets adds additional layer of robustness.

### Conclusions

CD8-IM demonstrated a moderately stronger association with CRC survival compared to other assessed single immune markers and immune cell score, both in two-tiered and three-tiered survival models, supporting the use of a single immune cell marker in prognostic stratification of patients with localized resected CRC. CD8-IM alone or in combination with effector molecules could serve as a predictive biomarker for adjuvant chemotherapy benefit in localized CRC or response to immune checkpoint inhibitors in advanced CRC in future prospective studies. Confirmation of these findings in an independent CRC cohort could significantly advance the integration of TILs into clinical practice, independent of reliance on commercial digital pathology platforms.

## Data Availability

Individual patient data and related tumor information underlying this article cannot be shared publicly due to data privacy protection laws. However, grouped data will be shared on reasonable request to the corresponding author.

## Contributors

DW and MH conceptualized the study. DW, MK, HB, and MH were responsible for data collection. DW, DE, and MH was responsible for data analysis and generation of figures. DW, DE, and MH were responsible for data interpretation. All authors verified the data. First version of the manuscript was drafted by DW. All authors participated in reviewing of the manuscript, approved the final draft for submission, have access to the reported data, and accept responsibility for the decision to submit for publication.

